# Adversarial Learning for MRI Reconstruction and Classification of Cognitively Impaired Individuals

**DOI:** 10.1101/2023.11.13.23298477

**Authors:** Xiao Zhou, Akshara R. Balachandra, Michael F. Romano, Sang P. Chin, Rhoda Au, Vijaya B. Kolachalama

## Abstract

Game theory-inspired deep learning using a generative adversarial network provides an environment to competitively interact and accomplish a goal. In the context of medical imaging, most work has focused on achieving single tasks such as improving image resolution, segmenting images, and correcting motion artifacts. We present a dual-objective adversarial learning framework that simultaneously (1) reconstructs higher quality brain magnetic resonance images (MRIs) that (2) retain disease-specific imaging features critical for predicting progression from mild cognitive impairment (MCI) to Alzheimer’s disease (AD). We obtained 3-Tesla, T1-weighted brain MRIs of participants from the Alzheimer’s Disease Neuroimaging Initiative (ADNI, N=342) and the National Alzheimer’s Coordinating Center (NACC, N=190) datasets. We simulated MRIs with missing data by removing 50% of sagittal slices from the original scans (i.e., diced scans). The generator was trained to reconstruct brain MRIs using the diced scans as input. We introduced a classifier into the GAN architecture to discriminate between stable (i.e., sMCI) and progressive MCI (i.e., pMCI) based on the generated images to facilitate encoding of AD-related information during reconstruction. The framework was trained using ADNI data and externally validated on NACC data. In the NACC cohort, generated images had better image quality than the diced scans (SSIM: 0.553 ± 0.116 versus 0.348 ± 0.108). Furthermore, a classifier utilizing the generated images distinguished pMCI from sMCI more accurately than with the diced scans (F1-score: 0.634 ± 0.019 versus 0.573 ± 0.028). Competitive deep learning has potential to facilitate disease-oriented image reconstruction in those at risk of developing Alzheimer’s disease.

## 1 Introduction

Detection of preclinical Alzheimer’s disease (AD) is advancing greatly through novel developments in structural (e.g., MRI) and functional (e.g., PET) neuroimaging [1]. The quality of the MRI scan, determined largely by the scanner itself, can play an important role in accurately representing regions affected by disease [2]. Since raw MRI scans are acquired in k-space, image reconstruction algorithms are needed to transform the acquired raw data into interpretable image representations. MRI reconstruction may involve several signal processing steps such as noise pre-whitening for phased array data acquisition, interpolation, filtering, and k-space to image space transformation [3]. Since all these steps are executed on the raw MRI data, the reconstruction process is not necessarily aimed towards accurate disease detection. Modifying the objective of reconstruction to ensure the generated images are of high quality while facilitating accurate diagnosis of disease can have profound practical implications.

We contend that a generative adversarial network (GAN) is ideally suited to accomplish such a task by leveraging the principles of the two-player zero-sum game from game theory to capture the distribution of brain MRIs that show evidence of mild cognitive impairment due to Alzheimer’s dementia pathology. GANs utilize the minimax principle from game theory, where the generator and the discriminator compete with each other to achieve a Nash equilibrium [4]. In the context of medical imaging, a classical GAN contains two players: 1) the generator, which takes images from one domain (e.g., low-quality brain MRIs) as input and attempts to create images similar to real training data (e.g., high-quality brain MRIs) and 2) the discriminator, which tries to distinguish the generator’s output from the real training data. The discriminator mainly serves to ensure that the generated MRIs adhere to a distribution of real brain MRIs.

### 1.1 Related work

GAN-based approaches have been successful in MRI reconstruction. Yang *et al*. developed a conditional GAN-based model for fast compressed sensing MRI reconstruction [5], resulting in output images with preserved perceptual image details. In a related study, Quan *et al*. developed a GAN framework with deeper generator and discriminator networks and cyclic data consistency loss for improved interpolation in the given undersampled k-space data, followed by the use of a chained network to improve the quality of image reconstruction [6]. Additionally, Shaul *et al*. proposed a two-stage GAN framework for MRI reconstruction from undersampled k-space data, which estimated the missing k-space samples and fixed aliasing artifacts in the image space [7]. Chen and collaborators developed a multi-level densely connected super-resolution framework that incorporated GAN loss to enhance low resolution T1-weighted MRI volumes [8]. Others have developed GAN-based deep learning frameworks to enable reconstruction of high-quality MRI scans from low quality scans with fast acquisition time [9–11].

Most of these efforts focused exclusively on accurate reconstruction as the sole objective, where the frameworks were optimized for producing higher quality images and/or for time-efficiency. Further, there is limited literature on the use of MRI reconstruction techniques in the setting of neurodegenerative diseases such as Alzheimer’s disease. Iglesias *et al*. recently published a public AI tool, SynthSR, that creates high quality isotropic T1-weighted brain MRIs using any clinical brain input scan [12]. Though they validated this tool with brains from persons with Alzheimer’s disease, the main objective of their tool was not for enhancing diagnostic ability, but rather for use in 3D morphometry software. To try to fill this gap in the literature, we recently developed a GAN-based approach that aimed to simultaneously meet two objectives [13]: 1) to generate higher quality MR images from 1.5T scans and 2) to improve AD classification performance when these generated images are used as input. While this study established a proof-of-principle that competitive deep learning can be leveraged to achieve multiple objectives, the cases that were utilized to achieve the goal (i.e., scans of multiple magnetic field strengths obtained on the same individuals taken at the same time) are not routinely available in most real-world clinical scenarios. Therefore, GAN approaches that can simultaneously address multiple objectives in a more practical setting can be tremendously helpful in the clinical setting, aiding physicians in prognostication of MCI earlier in the disease course without the use of invasive testing (i.e., lumbar puncture), since not all patients with MCI progress to AD [14].

Broadly, MCI is clinically diagnosed when patients show cognitive deficits on cognitive testing while still being able to carry out their instrumental activities of daily living (IADLs; e.g., cooking, managing finances, etc.). Once patients with MCI are unable to do any one IADL, they have clinically progressed to dementia (i.e., they had progressive MCI [pMCI]). Although medial temporal atrophy affecting the hippocampus is usually seen in MCI due to AD pathology and greater atrophy portends a worse prognosis, it is often quite difficult to ascertain hippocampal atrophy by visual inspection or with measures of cortical thickness early in the disease course (i.e., when patients are still in the MCI stage), as the imaging changes can be very subtle [15, 16]. This problem can be compounded when MRIs are of lower quality in resource-limited healthcare settings. In related AD research, classifying between pMCI and stable MCI (sMCI; i.e., patients with MCI who do not progress to dementia) is generally considered to be challenging as the structural differences in the brain are subtle [17–19].

In this work, we aim to train a dual-objective GAN to (1) reconstruct higher quality brain MRIs that also (2) accurately retain disease-specific features critical for prognostication of AD progression. We hope that this setting, due to its inherent challenge, could further demonstrate GAN’s potential in clinical utilizations. To achieve these objectives, we extend the aforementioned classical GAN architecture by introducing a classifier that attempts to distinguish the reconstructed brain MRIs from the generator as belonging to a subject with pMCI versus sMCI, thus encouraging the generator to create realistic brain images while retaining AD-related information. The classifier can also help to mitigate the feature hallucination problem, where generative networks can sometimes add or remove key elements from images [20]. We aim to implement this framework at the stage when images are already constructed from the raw MRI data. The practical implication is that existing national and international working groups with access to large imaging studies can obtain additional disease-driven insights without having to collect additional data.

### 1.2 Contributions

The main contributions of this paper are summarized below:

- We present a dual-objective adversarial learning framework that was able to reconstruct higher quality brain MRIs that retained disease-related information in the setting of MCI due to AD pathology. We show that including a classifier in the GAN architecture increases the predictive value of the generated images, likely by encoding more disease-related information in the reconstructed scans.
- Our framework was able to address stability problems commonly encountered in training GANs by balancing the learning speeds of the generator, discriminator, and classifier with fine-tuned learning rates and having additional training iterations for the generator and classifier.
- We validated the predictive value of the generated scans in distinguishing pMCI from sMCI to address potential concerns of feature hallucination in the reconstructed images.
- We used T1-weighted 3T brain MRIs and corresponding clinical data from two independent datasets and demonstrated that our dual-objective GAN is generalizable. In summation, our framework shows promise in being utilized by existing national and international working groups with access to large imaging studies to obtain additional disease-driven insights without having to collect additional data.

## 2 Materials and methods

### 2.1 Study population

We obtained access to clinical and neuroimaging data from the Alzheimer’s Disease Neuroimaging Initiative (ADNI) and the National Alzheimer’s Coordinating Center (NACC) to perform this study.

ADNI is a multi-center project focused on curating clinical, imaging, genetic, and biochemical biomarkers for studying AD [21]. Our study utilized 3T T1-weighted MRIs that were obtained at a visit where the respective participant received a diagnosis of MCI and had cerebrospinal fluid data available. Where multiple such events were present for a single person, MRIs were refined by selecting the scan collected at the most recent date, and by utilizing fully sampled scans where possible. Participants were assigned diagnosis labels of progressive MCI if they progressed to AD within (0-36] months (rounded down to the nearest month) and stable MCI if they did not progress to AD within 36 months (Table 1).

**Table 1:** Study population.

| (a) ADNI<sup>a</sup> cohort |  |  |
| --- | --- | --- |
|  | pMCI<sup>b</sup> | sMCI<sup>b</sup> |
| N (% Male) | 116 (57.8) | 226 (57.1) |
| Age, years | 73.0 ± 7.2 (116) | 71.2 ± 7.3 (226) |
| MMSE<sup>b</sup> (N) | 27.1 ± 1.8 (116) | 28.2 ± 1.6 (226) |
| Years of educ. (N) | 15.9 ± 2.7 (116) | 16.4 ± 2.6 (226) |
| APOE4 (#0/1/2 - N) | 38/58/20 | 138/75/13 |
| (b) NACC<sup>a</sup> cohort |  |  |
| N (% Male) | 95 (48.4) | 95 (62.1) |
| Age, years | 75.9 ± 9.4 | 73.5 ± 10.2 |
| MMSE<sup>b</sup> (N) | 25.9 ± 2.5 (55) | 27.3 ± 2.0 (75) |
| Years of educ. (N) | 15.4 ± 2.9 (95) | 15.7 ± 2.4 (95) |
| APOE4 (#0/1/2 - N) | 38/33/12 | 58/31/4 |

Demographics summary of the participants from the ADNI and NACC datasets. Age, MMSE, and years of education are reported as means with the respective standard deviations.
<sup>a</sup>ADNI = Alzheimer’s Disease Neuroimaging Initiative, NACC = National Alzheimer’s Coordinating Center
<sup>b</sup>pMCI = progressive mild cognitive impairment, sMCI = stable mild cognitive impairment, MMSE = Mini Mental State Exam

NACC is a large database containing research data from Alzheimer’s Disease Research Centers across the United States. We utilized clinical and imaging data from a data freeze on December 12, 2020. For each subject, we first identified the visits at which they were diagnosed with amnestic or non-amnestic MCI. MRIs were chosen to minimize the time between a clinical diagnosis of MCI and the date of MRI acquisition. If the shortest time between an MCI clinical visit and MRI was longer than 6 months, the subject was excluded. Occasionally, participants had multiple MRIs that satisfied these criteria; in these cases, only one of these MRIs was utilized (Table 1).

### 2.2 Image processing

We pre-processed MRIs using SPM12 (Statistical Parametric Mapping, https://www.fil.ion.ucl.ac.uk/spm/software/spm12/) and MATLAB version 2020a. T1-weighted MRIs were bias-corrected, normalized into MNI space and skull-stripped. We adopted an approach to skull stripping similar to the one described by Mitchell et al [22] in which SPM12-derived probabilistic maps of GM, WM, and CSF were summed and thresholded at 0.2 to produce binary whole-brain masks. We performed quality checks of each individual binary mask to ensure skull stripping was performed correctly and 1 subject from the ADNI cohort was excluded due to poor pre-processing results. We opted to use the SPM12 pipeline for skull stripping as it has been shown to accurately estimate total intracranial volume when compared with expert manual estimation [23]. The binary masks were then applied to the normalized, bias-corrected MRIs. Additional details can be found in our previously published work [24]. Henceforth, these normalized, bias-corrected, and masked MRIs will be referred to as ‘original scans’. ‘Diced’ scans were created by randomly selecting half of the sagittal slices of the original scans and replacing their values with zeroes. These diced scans were used as input to the generator. A schematic of our image processing workflow is provided in Figure 1.

**Figure 1:**
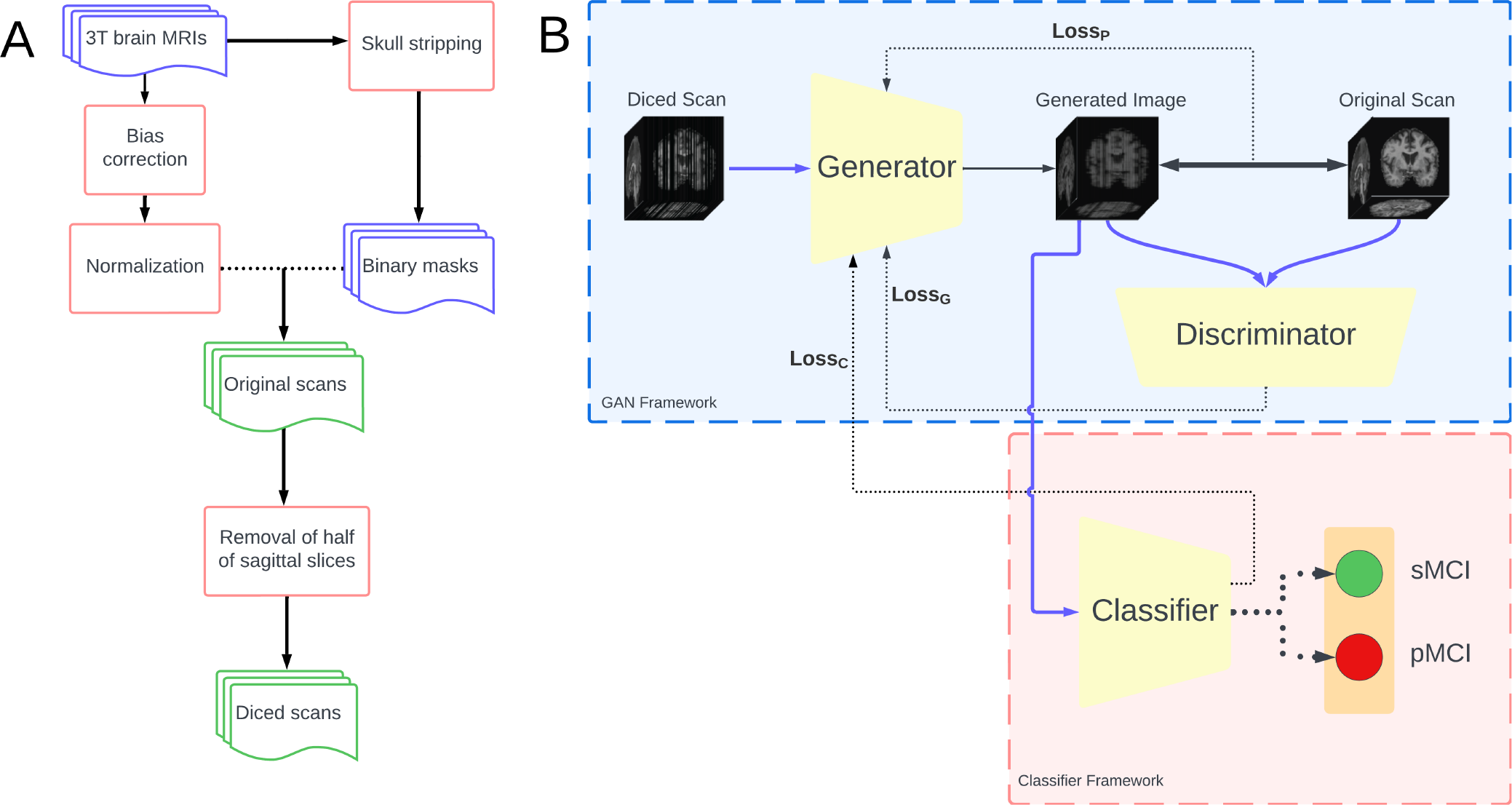
Image processing workflow and adversarial learning framework. (A) Raw T1 brain MRIs were pre-processed using SPM12 with bias-correction, normalization, and skull stripping. Binary whole-brain masks were applied to the normalized MRIs yielding the “original” images. Diced scans were created by randomly selecting half of the sagittal slices from the original scans and replacing them with zeroes. (B) Diced scans serve as input to the generator. The generator incorporates losses from the discriminator (Loss_G_), classifier (Loss_C_), and a new ‘perception loss’ (Loss_P_) derived from the comparison of the generated images with the corresponding reference images (i.e., Original scans). While the discriminator attempts to differentiate between the generated images and the original scans, the classifier uses the generated images to classify persons who have sMCI from those with pMCI. See Fig. 2 for a full schematic of the classifier.

For plotting Fig. 3, tissue probability maps obtained from SPM12 corresponding to CSF were thresholded at 0.2, and plotted as blue over the reconstructed images. A background mask was obtained by identifying relevant brain parenchyma as the sum of white matter, gray matter, and CSF tissue probability maps, thresholding at 0.2, and setting the background as the voxels not part of the brain parenchyma. These voxels were set to black in Fig. 3.

**Figure 2:**
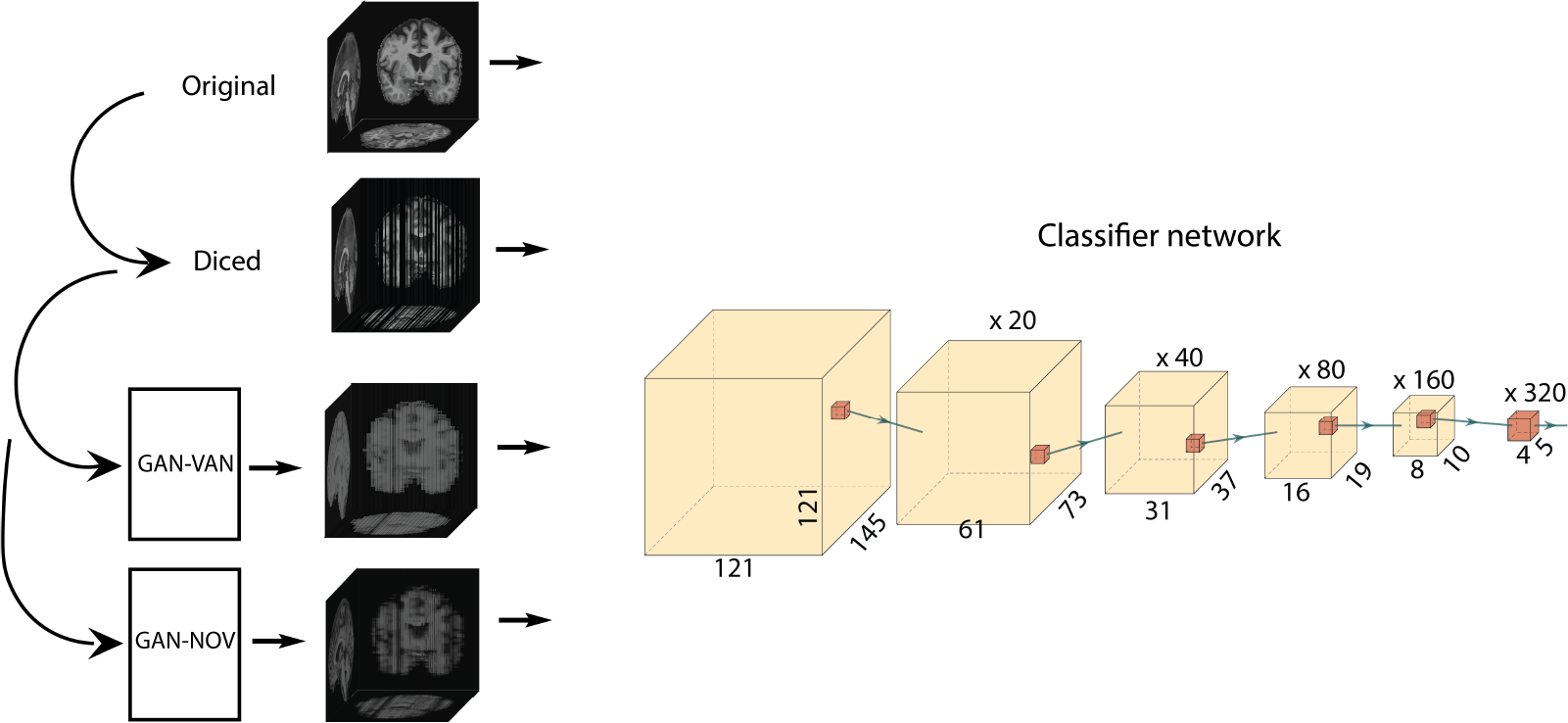
Schematics of the classifier. An exemplar MRI before (‘Original’) and after (‘Diced’) random removal of sagittal slices. Also shown are examples of brain MRIs following two different GAN-based methods of imputing the missing slices, GAN-VAN and GAN-NOV. On the right is a visual representation of the three-dimensional classifier network used to differentiate between stable and progressive MCI cases. The classifier network representation was generated using PlotNeuralNet v1.0.0.[25]

**Figure 3:**
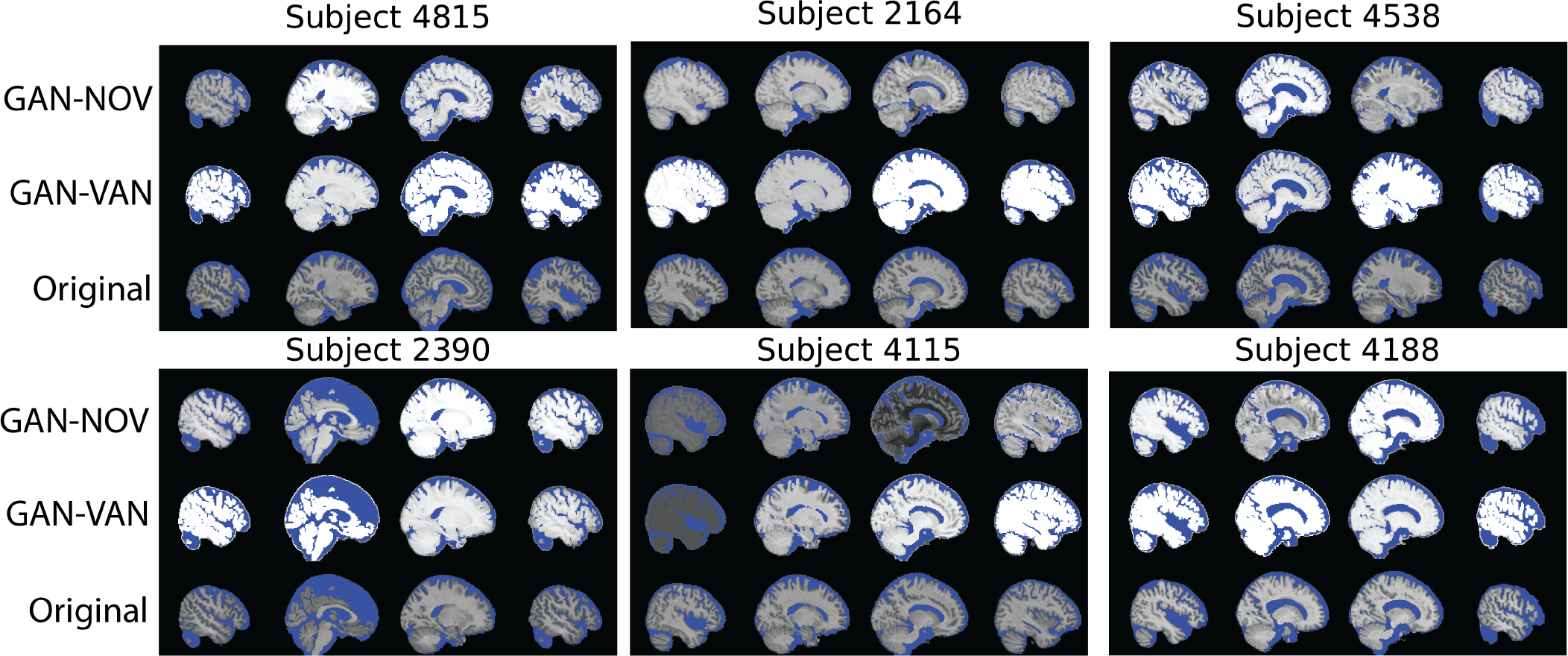
Sample reconstructed brain slices. Sample sagittal slices from six T1-weighted MRIs, labeled “Original” (bottom), and reconstructions, with signal intensities rescaled between the range [0, 1]. Reconstructed images are from either a generator utilizing perception loss, GAN loss, and classification loss together (GAN-NOV), or a generator utilizing perception loss and GAN loss only (GAN-VAN). Blue indicates CSF, while backgrounds were set to black.

### 2.3 Computational framework

We constructed an adversarial network for three-dimensional MR image reconstruction while simultaneously attempting to classify persons as having sMCI or pMCI. Our framework consisted of one generator, one discriminator, and one classifier. The generator takes diced scans as input and attempts to generate a more continuous version of the image. At the same time, the generated image is compared with the original scan via the discriminator. Concomitantly, the generated image is used as input to the classifier to predict if the original scan is from someone with sMCI or pMCI. Thus, our framework is performing two tasks, image reconstruction and classification. We incorporated three separate loss functions during model training, each corresponding to one part of our framework: 1) discriminator loss, 2) classification loss, and 3) perception loss (Fig.1). After the model was trained, we compared the generated images with the original scans using various image quality metrics. We also evaluated the predictive ability of the generated images by comparing the performance of a classifier trained on the generated images with a classifier trained on the original scans.

Let *Z* denote the set of input slices to be reconstructed, *z* as an instance of the set; *g* ∈ *G* as the reconstructed image, and *t* ∈ *T* as the original scan. To reconstruct a complete 3D volume given a few 2D slices, a model capable of generating missing slices based on spatial context is necessary. In an adversarial setting, the two networks are competing against each other and the objective can be formulated as

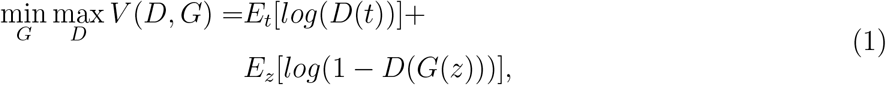

where *V* is the function value, *D*(*t*) is the discriminator’s output given *t* as the input, and *G*(*z*) is the generator’s output given *z* as input.

The objective of the discriminator is to distinguish between the generated and reference data. Since the output contains binary outcomes, binary cross entropy was used to measure the loss defined as

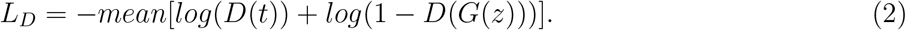

The objective of the generator is to create outputs that are indistinguishable from the real data. The loss for generator is defined as

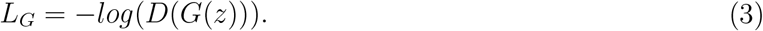

To guide the generator to reconstruct an image that captured disease-related information, we included binary cross entropy loss from the classifier, defined as

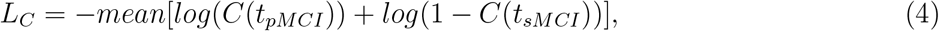

where *sMCI* indicates that the scan belongs to a person with sMCI, and *pMCI* indicates the scan is from a person with pMCI. We also introduced a perception loss that estimated the distance between the reconstructed image and the target image to restrict the generator from deviating from the target image, defined as

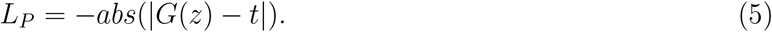

The complete objective function for the generator is defined as

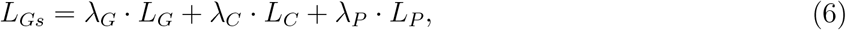

where *λ*_*G*_, *λ*_*C*_, and *λ*_*P*_ are the weights assigned for the corresponding losses.

Overall, the generator was composed of 2 convolutional and 2 transposed convolutional layers, where each of them have a kernel of size [3,1,1] with a padding of [1,0,0]. The convolutional layers had a stride of [2,1,1] to perform in lieu of discrete pooling layers. Batch normalization was applied for each layer’s output to rescale the parameters. The activation for the last layer was *tanh*, while the other layers used ReLU as activation function. The tanh activation function is used as the final layer of the generator because it has several benefits that helps the network have better results and greater training speed: (1) The tanh function outputs values in the range [−1,1], which is compatible with our pre-normalized MRI scan data. (2) It increases training stability and minimizes the risk of unbounded outputs by mapping extreme values into this bounded range (3) Due to its nature, the tanh function maps both positive and negative inputs away from zero, thereby promoting faster, yet smoother learning.

The discriminator was composed of 4 convolutional layers, where the first three had a kernel of size [3,3,3] with a padding of [1,1,1] and a stride of 3, and the last layer had a kernel of size [5,6,5] without padding and a stride of 1. Similar to the generator, we did not apply pooling layers for the discriminator, and batch normalization was applied to each layer’s output. We used LeakyReLU as the activation function for all layers except the last one. The last layer used the sigmoid activation function.

The classifier was composed of 6 convolutional layers, each with a doubled number of filters from the previous layer, starting from 20. The first five layers had a kernel of size [3,3,3] with a padding of [1,1,1] and a stride of 2. The last layer had a kernel of size [4,5,4] without padding and a stride of 1. We did not use any pooling layers. Batch normalization with a dropout rate of 0.25 was applied for each layer’s output. Similar to the discriminator, LeakyReLU was used as the activation function for all layers except the last one. The last layer used the sigmoid activation function.

The activation functions used by the discriminator and classifier were LeakyReLU in the hidden layers and sigmoid in the final layer to optimize for performance and stability. Sigmoid was chosen in the last layer because (1) it maps the output to the range [0, 1], which is suitable for our prediction tasks and (2) our early optimization efforts indicated that it performs best for our tasks. We chose not to use sigmoid in the hidden layers since (1) it has a small gradient during backpropagation, which may cause the gradients to vanish when overlapped multiple times, (2) it is more expensive computationally, which would make the training slower, and (3) its output range can make the learning complex, as it prevents negative correlations between neurons. Finally, LeakyReLU was selected in hidden layers because (1) it is more efficient than sigmoid and tanh, rendering faster training, and (2) it allows for a small gradient for negative outputs, which helps prevent the vanishing gradient problem. These choices are supported by previous work that used similar settings [13, 26, 27] and empirical results from our early optimization experiments.

Hyperparameter tuning was performed using Bayesian search [28], which was implemented using Gaussian Process (GP) to optimize the model and evaluate the relationship between the parameters and the performance. The formula used by the hyperparameter optimizer is

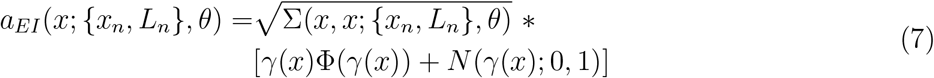

where

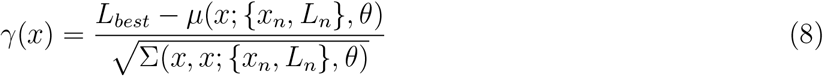

and

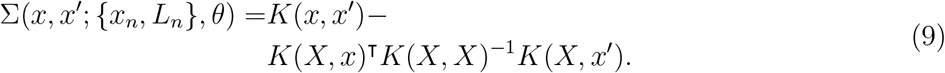

Note 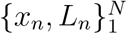 are the pairs of observations, and Φ is the cumulative distribution function of the standard Gaussian distribution (*N* (0, 1)), *γ*(*x*) is *Z*-score, *K*(*X, x*) is the *N* -dim vector of cross-co-variances between *x* and set *X* (which contains all *x*s). *K*(*X, X*) is the Gram matrix for set *X* (*K* is a kernel function), *m* : *χ* ⟶ *R* is the mean function, *θ* is the kernel parameters, and Matern 5*/*2 kernel was used (Table 2). We applied standard black-box optimization algorithms to optimize the function as described by Snoek *et al*. [29].

**Table 2:** Variable definitions.

| Variable | Description |
| --- | --- |
| $\{x_n, L_n\}_1^N$ | observation pairs |
| $\Phi$ | CDF of normal distribution |
| $\gamma(x)$ | $Z$-score |
| $K(X, x)$ | $N$-dim vector of cross-covariances of $x$ and $X$ |
| $K(X, X)$ | Gram matrix for set $X$ |
| $m : \chi \rightarrow R$ | mean function |
| $\theta$ | parameters of the kernel |

### 2.4 Statistics

We used Fisher exact tests to statistically compare categorical data and Mann-Whitney U tests to compare continuous variables. To compare proportions of persons with different numbers of APOE4 alleles between the ADNI and NACC cohorts, counts were pooled between bins. For example, to compare proportions of persons with 1 APOE4 allele between the two cohorts, the number of persons with 0 or 2 APOE4 alleles were combined.

#### Algorithm 1 GAN Model for 3D MR Image Reconstruction and Classification

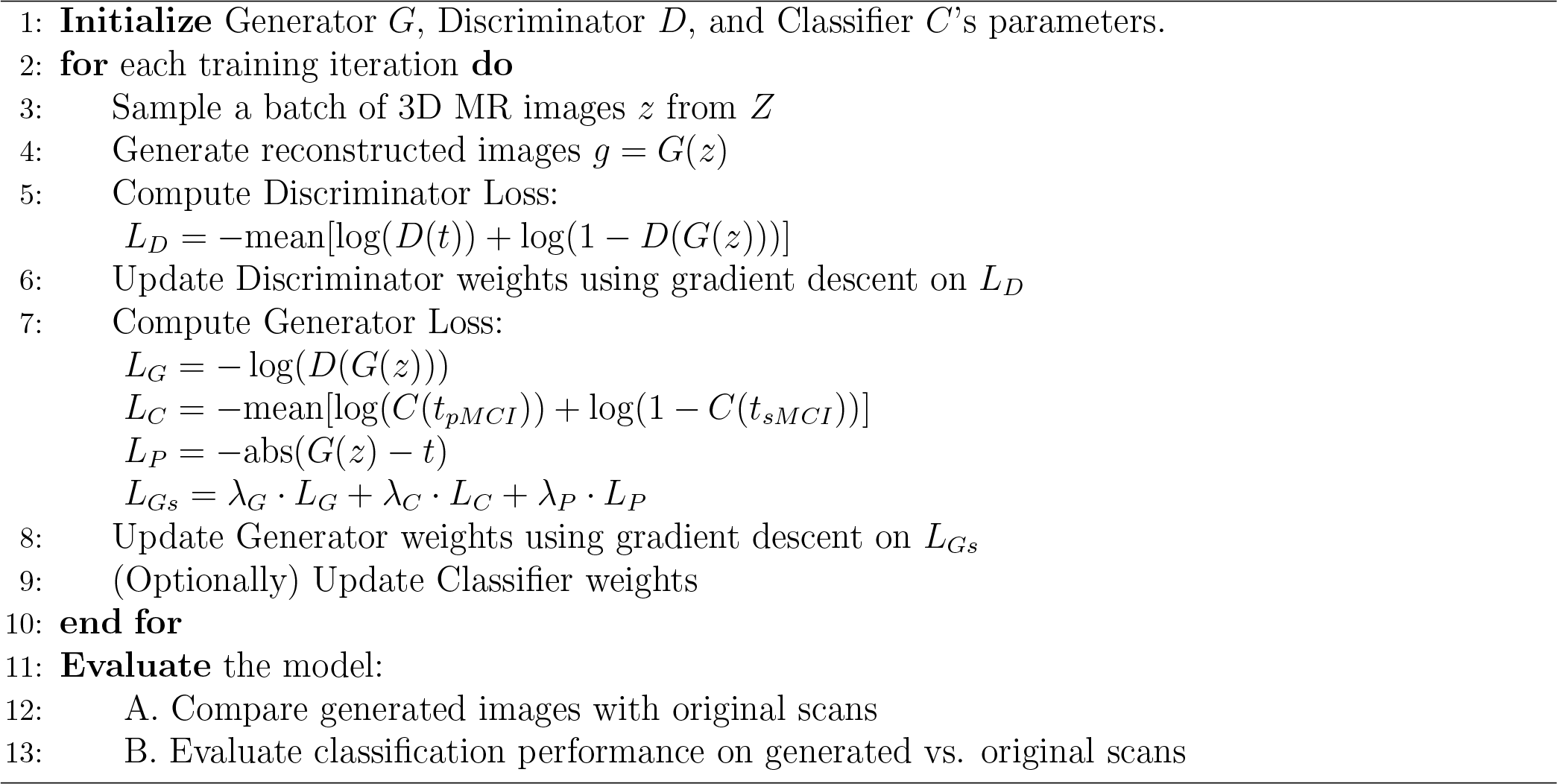

### 2.5 Data and code availability

Both ADNI and NACC datasets are publicly available and can be obtained directly from the respective sources. Python scripts and manuals are made available on GitHub (https://github.com/vkola-lab/jbhi2023)

### 2.6 Complexity Analysis

#### Time Complexity

In general, for convolutional operations, the time complexity is given by: *O*(*K*^3^ × *F* × *W* × *H* × *D*), where *K* is the kernel size, *F* is the number of filters, and *W, H, D* are the dimensions of the inputs.

In our case, for the discriminator and classifier, the approximate time complexity can be represented as *O*(3^3^ × 20 × 121 × 145 × 121). For the generator, the time complexity is approximately: *O*(3^3^ × 3 × 121 × 145 × 121).

#### Space Complexity

In our model, the space complexity of the network is mostly determined by the number of parameters in the convolutional layers, which in general is *O*(*K*^3^ × *F* × *C*), where *C* is the number of input channels.

For the discriminator and classifier, the approximate space complexity can be represented as *O*(3^3^ × 20 × 1). For the generator, the space complexity is approximately: *O*(3^3^ × 3 × 1).

#### Number of Parameters

The total number of parameters in our generative adversarial framework is the sum of the parameters for the generator, discriminator, and classifier, which is 1,864,189.

#### FLOPs (Floating Point Operations)

Our framework’s FLOPs are determined by the number of floating-point operations required during a forward pass. The FLOPs for the generator is 1,673,021,020, for the discriminator is 2,661,531, and for the classifier is 1,696,252,460.

## 3 Experiments

The ADNI dataset was randomly split 80% for training, 10% for validation, and 10% for internal testing. The optimal model was saved based on performance on the validation partition. The entire NACC dataset was used for external testing.

We trained the generator and the discriminator for 20 epochs with the Adam optimizer. The classifier was trained for 60 epochs with the stochastic gradient descent optimizer. Learning rates ranged from 0.001 to 0.005 for different components of the learning framework. Henceforth, we refer to this framework as GAN-NOV.

To evaluate the added benefit of the classification loss, we constructed another GAN-based reconstruction framework similar to the one described in Fig. 1, except now without a classifier being trained simultaneously. Thus, this generator only incorporates the GAN loss and perception loss. We refer to this network as GAN-VAN. GAN-VAN was trained in an identical fashion as GAN-NOV and used for comparison against GAN-NOV.

We generated images with GAN-VAN and GAN-NOV utilizing as input diced scans from the ADNI test partition and and NACC for testing. We computed various metrics to assess the image quality of the generated images.

To assess if our reconstruction frameworks were able to generate images that retained disease-relevant information, we trained and tested four separate CNN classifiers to discriminate between pMCI and sMCI utilizing unique sets of MRIs as inputs: original scans only, diced scans only, generated images from GAN-VAN only, and generated images from GAN-NOV only. The classifiers were initialized identically to the one in GAN-NOV and trained for 60 epochs with the stochastic gradient descent optimizer. The classifiers were trained, validated, and tested 5 times, each time with a different seed for the random splits of the ADNI cohort. We assessed the classification performance on the ADNI test partitions and on NACC.

### 3.1 Computing infrastructure

Model training and evaluation was performed on a GPU workstation with an NVIDIA 2080Ti graphics card containing 11 GB memory. We used PyTorch (v1.13.0) to implement the model. The training speed was about 10-15 min for each epoch, or about 0.12-0.19 iterations/s, and training took 3.5-4 hours to reach convergence. The inference speed was 1.17s per MRI scan with a batch size of 1.

### 3.2 Performance metrics

The image quality metrics contrast-to-noise ratio (CNR), signal-to-noise ratio (SNR), and structural similarity index measure (SSIM) [30] were used to compare the differences in quality between original scans and generated images. Two no-reference algorithms were also used for comparisons: Blind/Referenceless Image Spatial Quality Evaluator (BRISQUE) [31] and Perception-based Image Quality Evaluator (PIQE) [32]. Image quality metrics were calculated for each image in the ADNI test partition and the entire NACC cohort. We report the means and standard deviations of each metric for scans of the corresponding datasets in Table 3.

**Table 3:** Image quality results.

| (a). Results on ADNI test partition, batch normalization |  |  |  |  |  |
| --- | --- | --- | --- | --- | --- |
| Input Images | CNR | SNR | SSIM | BRISQUE | PIQE |
| Original | 2.111±0.274 | 0.686±0.052 | - | 43.513±1.594 | 41.936±2.062 |
| Diced | 0.726±0.131 | 0.537±0.046 | 0.319±0.065 | 43.567±0.333 | 78.307±5.322 |
| GAN-VAN | **2.235±0.299** | **1.857±0.214** | 0.544±0.051 | 42.485±0.741 | 72.028±3.068 |
| GAN-NOV | 1.934±0.335 | 1.437±0.395 | **0.580±0.059** | **42.246±0.563** | **68.120±1.351** |
| (b). Results on NACC cohort, batch normalization |  |  |  |  |  |
| Original | 2.065±0.294 | 0.676±0.074 | - | 44.292±2.940 | 43.182±6.298 |
| Diced | 0.696±0.156 | 0.526±0.074 | 0.348±0.108 | 43.742±1.256 | 78.442±4.695 |
| GAN-VAN | **2.193±0.359** | **1.821±0.230** | 0.523±0.105 | 42.563±1.128 | 72.133±4.024 |
| GAN-NOV | 1.790±0.363 | 1.432±0.534 | **0.553±0.116** | **42.505±1.066** | **68.176±2.079** |

Assessment of the classifiers was done by first generating receiver operating characteristic (ROC) and precision-recall (PR) curves based on model predictions on both the ADNI test partition and the NACC cohort. Then, for each ROC and PR curve, we computed the area under the curve (AUC) statistic. We computed the mean AUCs and standard deviations for each model across the 5 folds. We report mean values, along with the respective standard deviations, of model accuracy, precision, F1-score and Matthews correlation coefficient (MCC) in Table 4.

**Table 4:** Classifier performance.

| (a). Results on ADNI cohort test partition, batch normalization |  |  |  |  |
| --- | --- | --- | --- | --- |
| Input Images | Accuracy | Precision | F1-score | MCC |
| Original | 0.794±0.033 | 0.795±0.032 | 0.785±0.036 | 0.516±0.081 |
| Diced | 0.703±0.064 | 0.681±0.082 | 0.665±0.080 | 0.254±0.145 |
| GAN-VAN | **0.709±0.069** | **0.711±0.084** | 0.653±0.079 | 0.270±0.130 |
| GAN-NOV | **0.709±0.042** | 0.697±0.062 | **0.694±0.060** | **0.303±0.133** |
| (b). Results on NACC cohort, batch normalization |  |  |  |  |
| Original | 0.675±0.006 | 0.683±0.004 | 0.671±0.007 | 0.358±0.010 |
| Diced | 0.597±0.021 | 0.624±0.021 | 0.573±0.028 | 0.219±0.042 |
| GAN-VAN | 0.589±0.025 | 0.625±0.021 | 0.556±0.039 | 0.211±0.047 |
| GAN-NOV | **0.640±0.017** | **0.650±0.014** | **0.634±0.019** | **0.289±0.030** |

Means and standard deviation of contrast to noise ratio (CNR), signal to noise ratio (SNR), structural similarity (SSIM), blind/referenceless image spatial quality evaluator (BRISQUE), and perception-based image quality evaluator (PIQE) of all the corresponding images in the ADNI test partition and in NACC are shown. The set of best performing images in each metric is highlighted in bold text. Higher scores of CNR, SNR, and SSIM indicate better quality, while lower scores of BRISQUE and PIQE indicate better quality.

Performance of CNN classifiers on ADNI (a) and NACC (b). Performance values of classifiers utilizing either the original scans (**Original**), diced scans (**Diced**), or reconstructed images from **GAN-VAN** and **GAN-NOV** are shown. All CNNs were trained with the same parameters on 5 random splits of the ADNI dataset. Weighted averages for accuracy, precision, F1-score, and Matthews correlation coefficient (MCC) were calculated. The input images leading to the best performance when compared to the original scans in each metric within each cohort is highlighted in bold text.

## 4 Results

The primary objective of our study was to evaluate the efficacy of our multi-objective adversarial deep learning framework in MRI reconstruction and classification. We aimed to determine if our approach could produce high-quality images while retaining disease-relevant information. Our study populations did not differ in terms of sex or APOE scores in either the pMCI or sMCI groups (APOE: sMCI, all *p* = 1; pMCI, *p* = 0.23 (0 APOE4 alleles), *p* = 0.58 (1 APOE4 allele), and *p* = 1 (2 APOE4 alleles) (Bonferroni-corrected); sex: sMCI, *p* = 0.46; pMCI, *p* = 0.21). For sMCI, persons in the NACC cohort were older (*U* = 12852, *p* = 0.0053), had lower MMSE scores (*U* = 5862, *p* = 4.4*e* − 05, with 20 missing samples from NACC), and fewer years of education (*U* = 9112.5, *p* = 0.03). For pMCI, education did not differ significantly between the two cohorts (*U* = 4858, *p* = 0.13), though persons in the NACC cohort were older (*U* = 6699, *p* = 0.007) and had lower MMSE scores (*U* = 2356, *p* = 0.0054, 40 missing samples from NACC). There was a larger proportion of persons with at least 1 APOE4 allele in the pMCI groups in both NACC and ADNI (*p* = 0.034 (NACC) and *p <* 0.0001 (ADNI)). Details of these performance are provided in Fig. 4, Table 3 and 4, as discussed in following sections.

**Figure 4:**
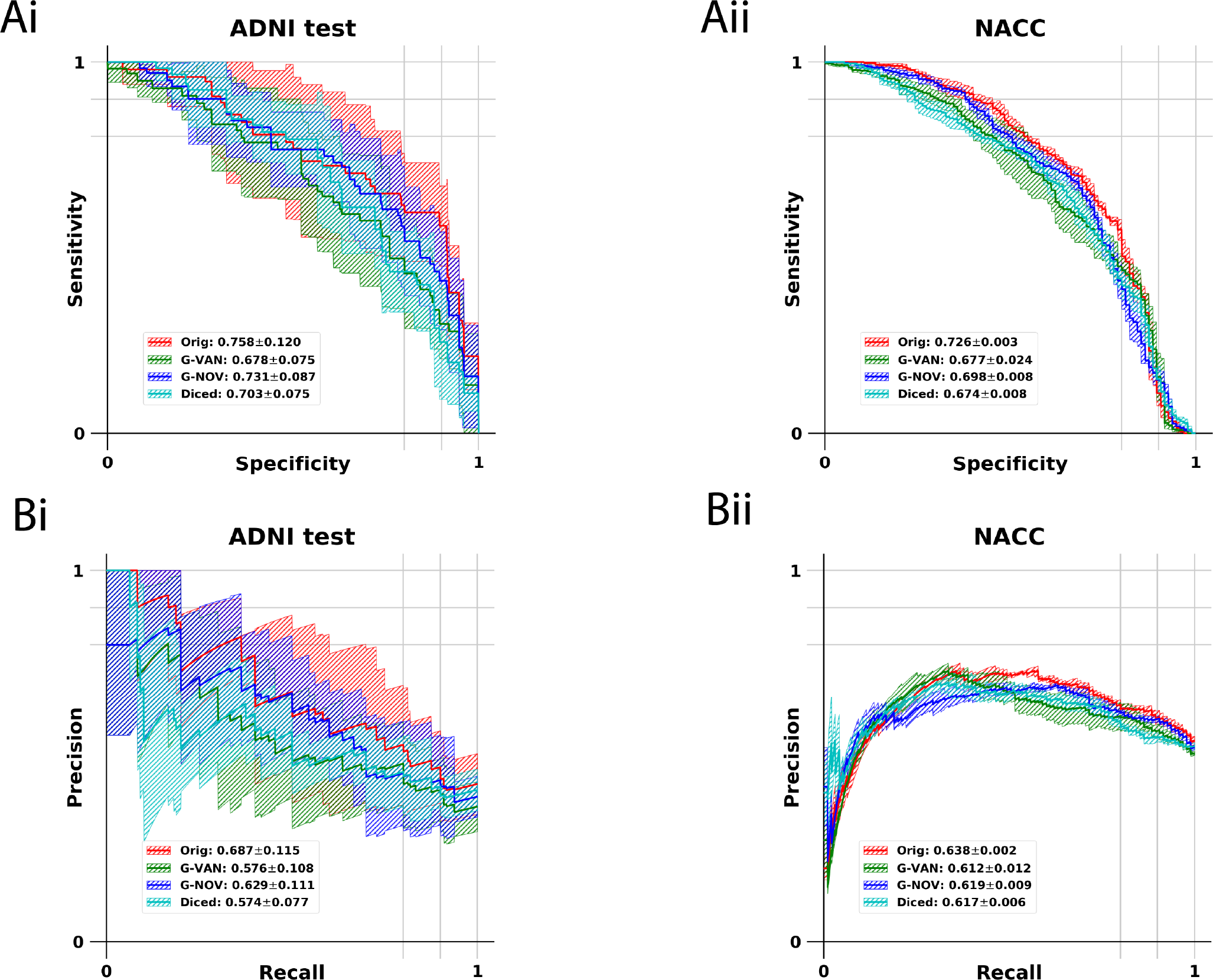
Performance of the CNN classifiers. Sensitivity-specificity (Ai and Aii) and precision-recall (Bi and Bii) curves comparing the four CNN classifiers on the ADNI test partition and NACC cohorts are shown. The classifier denoted as “Orig” was trained using original scans, “G-VAN” using generated images from GAN-VAN, “G-NOV” using generated images from GAN-NOV, and “Diced” using diced scans.

### 4.1 Qualitative assessment

To visually assess the quality of reconstructed images, we compared them with the original scans. This qualitative assessment is crucial as it provides an intuitive understanding of the reconstruction quality, which can be relevant for clinical applications. In Fig. 3, we show reconstructed missing sagittal brain slices of six sample MRIs using each reconstruction method and compare them with the corresponding original scan slice. We see that GAN-NOV captures the sulci and ventricles better than GAN-VAN. Additionally, cortical thinning and boundaries between the cortex and white matter appear more clearly on the images generated using the GAN-NOV framework. Different patterns of gray matter morphometry, particularly within the temporal and parietal lobes, can be seen in persons with MCI or AD, and it is therefore important to preserve these details for MCI and AD assessment [33]. Moreover, the GAN-VAN approach produces images that appear to have higher signal intensity and saturation of extreme values, whereas the images generated via the GAN-NOV framework have a more dynamic range of voxels.

### 4.2 Quantitative assessment

Various image quality metrics, which included some no-reference algorithms, were used to objectively compare the quality of the images produced by the GAN-based reconstruction methods with the original and diced scans. Overall, the GAN-based reconstruction techniques were able to generate images with improved quality (Table 3). As expected, the diced scans demonstrated the worst image quality across all metrics tested. Moreover, the images generated via the GAN-NOV framework outperformed those generated via GAN-VAN as evaluated using SSIM, BRISQUE, and PIQE. However, generated images from the GAN-VAN framework had improved CNR and SNR than those generated from the GAN-NOV framework. Interestingly, both GAN-VAN and GAN-NOV generated images with better BRISQUE scores than the original images (e.g., GAN-NOV, 42.505 versus Original, 44.292 on NACC).

To assess whether our framework was able to reconstruct the MR images while retaining disease-relevant information, we constructed CNN classifiers utilizing either the original scans, diced scans, or reconstructed images from GAN-VAN and GAN-NOV as input to discriminate between sMCI and pMCI cases. We trained the CNNs on 5 randomly selected folds of the ADNI dataset and performed internal testing on the corresponding ADNI test partitions and external validation on the NACC cohort. Performance of each classifier using various metrics is summarized in Table 4. ROC and PR curves for each classifier are presented in Fig. 4. As expected, the CNN classifier utilizing the original scans performed the best on both the ADNI test partition and NACC. Generated images from the GAN-NOV framework seemed to afford better discriminative ability than the diced scans (e.g., GAN-NOV, 64% accuracy versus Diced, 59.7% accuracy on NACC). While images generated by GAN-VAN generally demonstrated the highest CNR and SNR on both ADNI and NACC (Table 3), the classifier utilizing these images tended to perform similarly to, or in some metrics worse than, the classifier utilizing the diced scans on both datasets.

Images generated by GAN-NOV performed better than those from GAN-VAN across all reported metrics on the external testing dataset (i.e., NACC). A similar trend can be seen in Fig. 4, with the classifier using GAN-VAN-generated images performing similarly to the one using diced scans (AUC 0.677, GAN-VAN versus 0.674, Diced on NACC) and the classifier using images from GAN-NOV performing better with an AUC of 0.698. Overall, these results suggest that a generative adversarial framework utilizing an additional classification loss is able to reconstruct images of better quality while maintaining disease-relevant information and also appears to be generalizable to more heterogeneous datasets.

#### 4.2.1 Comparison of classifier performance

Discriminating between pMCI and sMCI is a well-known clinical challenge, as the pathological and imaging differences between these two conditions are more subtle than those between AD and normal cognition [17]. Further, predicting which individuals with MCI will progress to Alzheimer’s disease becomes more difficult the longer the lead time to clinical progression, especially when trying to predict 3 or more years prior to progression [34]. In light of these challenges, others have been able to predict which persons with MCI progress to AD with varying accuracy, between 0.5-0.8, using the widely available ADNI dataset [35–39]. Lian *et al*. were able to achieve one of the highest accuracies of 81% when classifying between pMCI and sMCI on the ADNI dataset [39], similar to our baseline classifier performance using the original scans. We achieved classification accuracy comparable to published studies of 64% with the generated images from GAN-NOV when tested on a completely independent dataset (i.e., NACC). This highlights some of our framework’s biggest strengths; we show GAN-NOV generates images that retain AD-relevant information essential for classification (i.e., diagnosis), and also that GAN-NOV is generalizable to unseen, heterogeneous datasets.

## 5 Discussion

In this work, we developed and validated a multi-objective adversarial deep learning framework for MRI reconstruction and classification of persons with sMCI and pMCI. We extend the classical GAN architecture by incorporating a classifier to reconstruct higher quality MR images with attention to disease-related information such that the reconstructed images can be used for prognostic purposes. We systematically trained and tested our MRI reconstruction framework using two independent datasets (ADNI & NACC), and presented the performance of our framework using well-known image quality and classification metrics.

Medical imaging plays a pivotal role in the diagnosis and monitoring of neurodegenerative diseases like AD. Some of the biggest challenges in identifying patients with MCI who are at high risk of progressing to AD lie in the heterogeneity and wide spectrum of presentation of MCI. Namely, cognitive testing appears to be poorly sensitive in identifying patients with pMCI, with up to 50% of patients with pMCI endorsing no memory complaints 3 years prior to progression [40]. In the ADNI dataset, prior studies have found a significant proportion of subjects with MCI to have normal cognitive test results and no significant cortical atrophy, even though many of these subjects progressed to AD [14, 16]. Even though cortical atrophy can be subtle and cognitive testing may not show deficits in a sizable proportion of pMCI patients, our competitive deep learning framework generated images with greater AD-related information than the Diced scans, leading to improved performance in identifying persons with pMCI. One of the strengths of our framework is that we attempted to mitigate the feature hallucination problem [20] inherent to generative networks by validating the MRI reconstructions to ensure accurate encoding of disease-related information, which is a necessary prerequisite to eventual use of such a framework in the clinical setting.

Previous work has focused on MRI reconstruction using downsampled brain scans of healthy individuals [41–43] or of patients with other neurological illnesses like multiple sclerosis [7]. Although Eo *et al*. proposed a deep-learning reconstruction framework in the AD population using scans from ADNI, they did not assess if the reconstructed images maintained AD-related information or validate their framework on an independent patient cohort [44]. Further, most of the published frameworks have tackled reconstruction and superresolution in the raw MRI k-space, which limits their applicability to large, established imaging databases for Alzheimer’s disease. Our model can effectively reconstruct high-quality MRIs from scans with missing slices, ensuring vital disease-related features are retained for accurate pMCI and sMCI differentiation. This advancement allows for quicker scanning protocols, reducing scan time and resource use. Extending to MRI super-resolution, our framework mirrors a GAN-based method [45], enhancing MRI detail by generating thinner slices from thicker ones and enriching information content. Such improvements in scan efficiency can increase patient comfort, lower healthcare costs, expand MRI accessibility, and minimize the risk of motion artifacts by shortening required stillness durations.

Our proposed framework utilizing the GAN loss, perception loss, and classification loss generated images that boosted classification accuracy from 59.7% (utilizing diced scans) to 64%. It appears that inclusion of perception loss without classification loss, like in GAN-VAN, did not yield a generalizable framework, as the classifier utilizing GAN-VAN-generated images performed worse in most metrics than the one using diced scans (Table 4). Images generated by GAN-NOV may encode more disease-related information due to inclusion of the classification loss, thus allowing the generator network to potentially learn latent imaging features that are weakly associated with disease across MRI scans in different datasets (i.e., ADNI and NACC). These latent features may be predictive of progression from MCI to AD, but may not directly contribute to image quality. Since GAN-VAN does not incorporate classification loss, it likely cannot learn these latent features when generating images and instead focuses more on features relating to image quality.

GAN-VAN generated images with the highest SNR and CNR, likely due to the greater average pixel intensities causing oversaturation and clipping, leading to simultaneously higher mean pixel values and decreased variance. Since SNR and CNR are directly proportional to mean pixel values and inversely proportional to the variance, these measures are likely artificially inflated. In contrast, GAN-NOV generated images that demonstrated better SSIM, suggesting these images have less structural distortion when compared with the original scans than those from GAN-VAN. It is important to note image quality assessment should be done using a variety of metrics, not just SNR and CNR, especially since these latter two measures are known to be inferior metrics when comparing distorted images against corresponding references, like we do in this paper [30, 46]. For such image quality analysis, SSIM appears to be superior [30].

We attempted to tackle multiple technical challenges that often arise when training GAN frameworks. For example, during earlier epochs of training, it is possible for the discriminator to dominate, as it is straightforward to distinguish the generated image from the original (i.e., target) image. To mitigate this issue, we tuned the learning rate and paused the discriminator periodically to prevent it from learning faster than the generator. Additionally, as discussed earlier, to ensure that the generator learns the appropriate way to reconstruct images with reference to the original scans, we introduced perception and classification losses as additional weighted loss terms.

### 5.1 Limitations

Our study has a few limitations. We restricted the network depth in various settings due to hardware limits (GPU memory), including the number of filters (both generator and discriminator), batch size, and number of convolutional layers. We hypothesize that a larger network architecture may significantly enhance the performance of the model. However, the high-quality scans for sMCI and pMCI brains are insufficient for training a high-quality generator, which generally requires thousands of samples. Though the generator is able to obtain reasonable predictions on the NACC dataset, its broader applicability needs further verification on other datasets. It remains a challenge to maintain the balance between the different components of competitive networks even with our modified training method. For example, while the discriminator could maintain stability during the early phases of training, it may prove to be ineffective in the late phase. In other words, as the generator improves, it tends to be rather difficult for the discriminator to identify the differences and similarities between the generated image and the reference image, especially when the generated image belongs to the reference distribution. To alleviate this issue, potential resolutions include (1) introducing additional/alternative loss to adjust the difference during training, (2) providing additional input as extra information (i.e., spatial information) to the discriminator in later phase, (3) or using additional discriminators to create an ensemble for higher performance in later training epochs.

### 5.2 Future directions

Sufficient spatial resolution is essential for identifying potentially minute structural brain changes that can occur in the early stages of MCI and AD. However, MRIs require long acquisition times, which can lead to patient discomfort and motion artifacts. Thus, it is essential to achieve an ideal balance between image quality and scan time. Parallel sequences can speed acquisition time, but compared to fully sampled sequences, have lower signal-to-noise-ratios [47]. Our framework could be used in 2D MRI to acquire fewer images while maintaining adequate resolution of the generated slices. Additionally, our framework could address issues such as zipper artifacts, which appear linearly in the image space [48]. Our framework could also be extended to perform slice imputation recursively, which could enable sparser sampling or improved resolution. For example, a generated slice could be used as training data to generate further slices. Lastly, since we designed our framework to work with brain MRIs curated from existing imaging databases, our method could be easily extended to other imaging data banks for conditions other than AD such as vascular dementia, epilepsy, or multiple sclerosis.

## 6 Conclusion

In conclusion, we proposed an adversarial learning framework that takes T1-weighted MRI scans with missing slices as input and generates images with improved quality while maintaining disease-related information, assessed by discriminating between progressive MCI from stable MCI using the generated images. We trained our framework on cases from ADNI and tested it on an independent, heterogeneous dataset, NACC. We note that this framework is able to meet dual objectives even when its learning ability is largely limited due to hardware and sample size. We found that the classification loss is crucial to guide the generator in producing images that retain disease-related information and to improve generalizability of the framework. Further validation of our framework, preferably using gold-standard evidence such as post-mortem neuropathology or PET imaging, is needed to confirm the utility of GAN-based frameworks for image reconstruction and cognitive assessment.

## Data Availability

All data produced in the present study are available upon reasonable request to the authors.

